# Accuracy of the drug dependency checkbox on the Maine birth certificate for Medicaid-covered births, 2016-2020

**DOI:** 10.1101/2022.07.14.22277138

**Authors:** Julia Dudley, Catherine McGuire, Apsara Kumarage, Chinonye Anumaka, Katherine A. Ahrens

**Affiliations:** Muskie School of Public Service, University of Southern Maine, Portland, ME; Cutler Institute, University of Southern Maine, Portland, ME

## Abstract

**Introduction:** The accuracy of the drug dependency checkbox on the Maine birth certificate is unknown. Our objective was to compare the drug dependency checkbox with information on substance use disorders as documented in Medicaid claims data.

**Methods:** Using rule-based deterministic matching, we linked Medicaid enrollment information to 2016–2020 Maine birth record data (N=58,584). Among the linked records (n=27,448), we identified maternal substance use disorder (SUD) diagnoses during the 280 days before through 7 days after delivery using ICD-CM-10 diagnosis codes. We used the following hierarchy to create mutually exclusive SUD categories: opioid use disorder (OUD), cannabis use disorder without cocaine use disorder, and other SUD disorders (alcohol, cocaine, nicotine, or other).

**Results:** Among women enrolled in Medicaid at the time of delivery, 12% had drug dependency indicated on their birth record and 33% had at least one SUD diagnosis recorded in their Medicaid claims. Among the birth records with the drug dependency indicated, 56% had an OUD, 25% cannabis use disorder without cocaine use disorder, 8% other SUD, and 10% had no SUD. Among those without drug dependency indicated, the corresponding percentages were 4%, 9%, 13%, and 75%.

**Discussion:** Although diagnoses of OUD and cannabis use disorder were more common among birth records with the drug dependency checkbox checked, reporting of drug dependency on the birth record does not appear to accurately indicate SUD during pregnancy.

**Conclusions:** Our findings suggest the drug dependency checkbox on the Maine birth certificate may be of limited value in identifying SUD during pregnancy.

## INTRODUCTION

Maine has one of the highest rates of maternal opioid use disorder (OUD) in the United States (US).^1^ In 2018, the prevalence of maternal OUD at delivery hospitalization was 34.9 per 1000 deliveries (nearly 3.5%) – a more than 40-fold increase since 1999.^2,3^ In addition, drug overdose mortality rates are at an all-time high in the US and Maine, with Maine’s drug overdose mortality rate nearly 50% higher than the nation overall (45.3 vs. 31.3 drug overdose deaths per 100,000 in 2021).^4^ Maine has the highest percentage of residents living in a rural area (61%)^5^ and rural populations are at higher risk of morbidity and mortality associated with opioid drug use.^6^ Together, these factors warrant renewed efforts to focus on decreasing the prevalence of OUD during pregnancy, and reducing the adverse health consequences of maternal OUD, such as opioid exposure *in utero*.

The MaineMOM (Maternal Opioid Misuse) program began in 2020 with funding from Centers for Medicare & Medicaid Services (CMS) of the U.S. Department of Health and Human Services. ^7^ The program aims to improve care for Maine’s low-income pregnant and postpartum women with OUD by working to increase access to high-quality care, while reducing care costs and better integrating substance abuse treatment and maternal care. As part of the MaineMOM evaluation, we linked birth records in Maine to Medicaid administrative data. Evaluation of care performance measures though birth record-Medicaid linkages has been done annually for more than 10 years;^7^ however, the MaineMOM evaluation provided an opportunity to do quarterly linkages and compare information captured between the two data sources. A drug dependency checkbox, added to the Maine birth record in 2013, had never been compared with information from other data sources on substance use disorder.

Therefore, the objective of our analysis was to estimate the accuracy of the drug dependency checkbox on the Maine birth record compared with information on substance use disorders as documented in the claims data for Medicaid-covered births.

## METHODS

### Data Sources

We used data from records for live births that occurred in Maine to Maine residents of all ages during 2016-2020, as provided by Data, Research and Vital Statistics (DRVS), Maine Center for Disease Control & Prevention, Maine Department of Health and Human Services (DHHS). These data were transferred to the Muskie School of Public Service using a secure file transfer portal. Medicaid enrollee information, medical claims, and pharmacy claims data from 2010 to 2020 were transferred securely to Muskie servers and updated monthly. Access to these restricted use data and permission to use these data for our program evaluation project were granted through a cooperative agreement between the University of Southern Maine and DHHS, a data use agreement between University of Southern Maine and DRVS, and a determination of non-human subjects research by the University of Southern Maine’s Institutional Review Board.

### Birth Record-Medicaid Linkage, Ever Enrolled in Medicaid

We linked Medicaid/Children’s Health Insurance Program (CHIP; hereafter together referred to as “Medicaid”) enrollment information to birth record data using rule-based deterministic matching, following a methodology described in a training provided by AcademyHealth, on behalf of the US Centers for Disease Control and Prevention^8^ and CMS, that was attended by University of Southern Maine staff (A.K.). Using fields available in both data sources, we constructed a linkage-algorithm consisting of 12 rules and ranked them in order of our confidence in yielding a true match (see **Supplemental Table 1, 3**). The fields used for matching included maternal date of birth (either full date or month and year only); name (various combinations of first name, first name first letter, middle name first letter, current last name, maiden name, child’s last name); Medicaid identification number (ID); and last 4 digits of Social Security Number. Medicaid ID and Social Security Number are collected on the Maine birth record; however, by Maine statute, the 9-digit Social Security Number can’t be used for purposes other than for administration of the Social Security Act plan.^9^

We then identified rows from the birth record data containing maternal information that was an exact match with information from the Medicaid enrollee data. Among the rule-based matches, we selected the match with the most discriminating combination of fields for each birth record. For the birth records that didn’t match, we identified those with Medicaid listed as the expected payer and/or there was a Medicaid ID number for the mother. For this group of birth records, we did matches manually, visually scanning Medicaid claims for a delivery that took place on the same day to a woman with the same name, birthdate, and/or Medicaid ID. Finally, we identified some maternal matches after their infants were linked to their birth record information (using a separate linkage algorithm). After each preliminary set of matches, we evaluated the quality of the matches by comparing information in the two data sources to make sure there were not obvious false linkages.

### Maternal Medicaid Enrollment at Time of Birth

For each match identified, we further determined if the woman giving birth was enrolled in full-benefit Medicaid coverage during the month the birth took place and if we could find her delivery-related claims. We identified full-benefit coverage using Recipient Aide Code(s) (RAC) documented for the delivery month in the Medicaid eligibility file. We scanned Medicaid medical claims for delivery-related claims to link to the birth episode using an algorithm and code list from SUPLN MODRN (State University Partners Learning Network, Medicaid Outcomes Distribute Research Network).^10,11^ We allowed for a 21-day window between the date of birth on the birth record and the date of delivery we identified in the claims data to provide leeway for dates not being a perfect match.

### Birth Record Maternal and Pregnancy Characteristics

Maternal and pregnancy characteristics were based on information collected in the birth record, as reported by the pregnant woman or transcribed from the medical record.^12,13^ Maine has used the 2003 version of the US Standard Certificate of Live Birth since 2013.^14^ Maternal characteristics included maternal age, race/ethnicity, education, and marital status. Pregnancy characteristics included plurality (singleton or plural birth), birthweight (categorized as low birthweight <2500 g), and gestational age at birth (categorized as preterm <37 completed weeks). We constructed a measure of facility volume, which was the average number of births per year at each birthing facility (categorized in 10-249, 250-499, 500-999, 1000-2499, and ≥2500 births).

### Revised-Graduated Prenatal Care Utilization Index (R-GINDEX)

Using information from the birth record, we calculated the revised-graduated prenatal care utilization index (R-GINDEX) to measure adequacy of prenatal care received during pregnancy.^15^ This index is based on the timing and number of prenatal visits recommended by the American College of Obstetricians an and Gynecologists and measures the usefulness or adequacy of care.^16^

### Birth Record Drug Dependency Checkbox

On August 1, 2013, a checkbox was added to the Maine state birth record to indicate “Alcohol or drug abuse". This checkbox was located within the section for “Risk factors for this pregnancy", which contained a series of checkboxes and the instructions “Check all that apply.” The drug dependency item was not part of the US Standard Certificate of Live Birth. No guidance was provided to the birth record certifier for the circumstances for which this checkbox should be checked.

### Substance Use Disorder Diagnosis During Pregnancy

Among women who were enrolled in Medicaid at the time of delivery, we scanned their medical claims to identify maternal substance use disorder diagnoses during the 280 days before through 7 days after delivery using ICD-CM 10 diagnosis codes related to substance use, abuse, dependence, and adverse effects (see footnote of **Supplemental Table 4** for code list).^17,18^

### Medication for Opioid Use Disorder During Pregnancy

Among women with at least one OUD diagnosis during pregnancy, we scanned their medical and pharmacy claims to identify any use of medication for their opioid use disorder (MOUD) during the 280 days before through 7 days after delivery. We looked for procedure codes indicating MOUD, such as ‘H0020’ for methadone administration. We also looked for National Drug Codes indicating a prescription was filled for buprenorphine, naltrexone (oral), injectable naltrexone, or buprenorphine/naloxone. We used a code list from a previously published multi-state Medicaid data analysis of MOUD prevalence.^19^

### Statistical Analysis

Among women enrolled in Medicaid at the time of delivery, we tabulated birth records by drug dependency checkbox status and if any substance use disorder (SUD) diagnosis was found in the Medicaid claims data. We also parsed out the SUD diagnoses by SUD type and, for those with OUD, by whether they received MOUD. We used the following hierarchy to create mutually exclusive SUD categories: OUD, cannabis use disorder without cocaine use disorder, and other SUD disorder (alcohol, cocaine, nicotine, or other substance use diagnosis). Our hierarchy prioritized OUD and combined less commonly encountered SUD into one category to avoid small cell counts. Per our data use agreement with DRVS, we suppressed presentation of results based on cell counts between 1 and 9.

### Sensitivity Analysis

We reran the main tabulations using only women with full benefit and non-dual enrollment during their entire pregnancy (for the 9 months preceding the delivery month). We did this to see how the findings would change if we restricted the analysis to women whose medical claims we could fully observe during pregnancy.

## RESULTS

We examined 58,584 live births in Maine from 2016-2020. Of the total births, 47% were to women enrolled in Medicaid at the month of delivery (**Table 1**; see **Supplemental Table 2** for results by birth year). Of the total births, 8% were to women who were enrolled in Medicaid at the month of delivery, but Medicaid was not reported as the primary payer on the birth record and 0.7% reported Medicaid as the primary payer but were not enrolled in Medicaid at the month of delivery. Separately, we found Medicaid delivery-related claims for 43% of total births.

**Table 1.** Medicaid linkage to Maine birth records for 58,584 in-state resident livebirths, 2016-2020

|  | Medicaid Enrolled at Month of Delivery |  | Medicaid Delivery-related Claim Found |  |
| --- | --- | --- | --- | --- |
|  | Yes | No | Yes | No |
| Primary Payer Reported on Birth Record |  |  |  |  |
| Medicaid | 22,837 (39.0) | 429 (0.7) | 22,359 (38.2) | 907 (1.5) |
| Non-Medicaid | 4,611 (7.9)* | 30,707 (52.4) | 3075 (5.2)* | 32,243 (55.1) |
| Total | 27,448 (46.9) | 31,136 (53.1) | 25,434 (43.4) | 33,150 (56.6) |

\*Approximately half of the women Medicaid Enrolled at Month of Delivery who reported their primary payer as non-Medicaid on the birth record were also enrolled in either Medicare (n=107) or private insurance (n=2160) at month of delivery (2267/4611). Approximately half of the women Medicaid Delivery-related Claim Found who reported their primary payer as non-Medicaid on the birth record were also enrolled in either Medicare (n=98) or private insurance (n=1298) at month of delivery (1396/3075).

Among the full sample of birth records, 13% of women with Medicaid listed as the primary payer had drug dependency indicated on the birth record, which was higher than that for women whose primary payer was reported as private insurance (2%), self-pay (6%), Tricare (2%), and other/unknown (4%), but lower than that for births whose primary payer was other government (e.g Medicare) (18%) (**Table 2**). Among the 27,448 birth records to women enrolled in Medicaid at the month of delivery, 12% had drug dependency indicated on their birth record and 33% had at least one SUD diagnosis recorded in their Medicaid claims during pregnancy.

**Table 2.** Medicaid linkage to Maine birth records for 58,584 in-state resident livebirths by drug dependency checkbox on birth record and substance use disorder diagnosis in Medicaid claims, 2016-2020

| Primary Payer Reported on Birth Record | Full Sample of Birth Record, N | Medicaid Enrolled at Month of Delivery, N (%) | Drug Dependency on Birth Record<br>N=58,584<br>Among Full Sample Birth Records |  | Drug Dependency on Birth Record<br>N=27,448<br>Among Medicaid Enrolled at Month of Delivery |  | Any Substance Use Disorder Diagnosis in Medicaid Claims*<br>N=27,448<br>Among Medicaid Enrolled at Month of Delivery |  |
| --- | --- | --- | --- | --- | --- | --- | --- | --- |
|  |  |  | Yes, n (%) | No, n (%) | Yes, n (%) | No, n (%) | Yes, n (%) | No, n (%) |
| Medicaid | 23,266 | 22,837 (98.2) | 2,954 (12.7) | 20,312 (87.3) | 2899 (12.7) | 19938 (87.3) | 8,389 (36.7) | 14,448 (63.3) |
| Private Insurance | 31,471 | 3,426 (10.9) | 466 (1.5) | 31,005 (98.5) | 198 (5.8) | 3228 (94.2) | 370 (10.8) | 3,056 (89.2) |
| Self-Pay | 2,488 | 862 (34.7) | 143 (5.8) | 2,345 (94.2) | 88 (10.2) | 774 (89.8) | 207 (24.0) | 655 (76.0) |
| Indian Health Service | 39 | 14 (35.9) | † | 35 (89.7) | † | 11 (78.6) | † | † |
| Champus/Tricare | 774 | 64 (8.3) | 15 (1.9) | 759 (98.1) | † | 55 (85.9) | 12 (18.8) | 52 (81.2) |
| Other Government | 133 | 67 (50.4) | 24 (18.1) | 109 (81.9) | 22 (32.8) | 45 (67.2) | 40 (59.7) | 27 (40.3) |
| Other/Unknown | 413 | 178 (43.1) | 18 (4.4) | 395 (95.6) | † | 169 (94.9) | 31 (17.4) | 147 (82.6) |
| Total | 58,584 | 27,448 (46.9) | 3,624 (6.2) | 54,963 (93.8) | 3228 (11.8) | 24220 (88.2) | 9,055 (33.0) | 18,393 (67.0) |

\*Substance use disorders include alcohol related disorders; opioid related disorders; cannabis related disorders; sedative, hypnotic, or anxiolytic related disorders; cocaine related disorders; other stimulant related disorders; hallucinogen related disorders; nicotine dependence; inhalant related disorders; and other psychoactive substance use related disorders.
†Data counts between 1 and 9 were suppressed for privacy purposes.

Several maternal and pregnancy characteristics varied by Medicaid enrollment at the month of delivery (**Table 3**). Characteristics more common among women enrolled in Medicaid were younger age, non-Hispanic White race/ethnicity, high school educational attainment, and being unmarried. R-GINDEX scores, birth weight, and rates of plurality and preterm births were similar to total births. Of the Medicaid-enrolled births, 56% occurred in facilities with <1000 deliveries annually, compared to 51% of total births.

**Table 3.** Maine birth records for 58,584 in-state resident livebirths by Medicaid enrollment at month of delivery, drug dependency checkbox, and hierarchical categorization of substance use disorder diagnosis in medical claims during pregnancy, 2016-2020

| Characteristics | Total Sample, %<br>(N = 58,584) | Medicaid Enrolled at Month of Delivery, %<br>(n= 27,448) | Drug Dependency = YES on Birth Record (n=3,228) |  |  |  | Drug Dependency = NO on Birth Record (n=24,220) |  |  |  |
| --- | --- | --- | --- | --- | --- | --- | --- | --- | --- | --- |
|  |  |  | Opioids, %<br>(n= 1,819) | Cannabis, %<br>(n= 821) | Alcohol, Cocaine, Nicotine, Other*, %<br>(n=251) | No SUD, %<br>(n= 337) | Opioids, %<br>(n= 872) | Cannabis, %<br>(n= 2,206) | Alcohol, Cocaine, Nicotine, or Other*, %<br>(n= 3,086) | No SUD %<br>(n= 18,056) |
| Total, row % |  |  | 56.4 | 25.4 | 7.7 | 10.4 | 3.6 | 9.1 | 12.7 | 74.5 |
| Maternal age, mean, y | 29.1 | 27.2 | 29.2 | 25.5 | 27.1 | 26.5 | 28.8 | 25.4 | 27.0 | 27.3 |
| Maternal age, y |  |  |  |  |  |  |  |  |  |  |
| <18 | 0.8 | 1.5 | † | 1.5 | † | † | † | 2.1 | 1.3 | 1.6 |
| 18-19 | 3.0 | 5.7 | 0.9 | 8.8 | 4.0 | 7.4 | 3.0 | 10.5 | 6.1 | 5.6 |
| 20-24 | 18.5 | 28.7 | 15.9 | 37.6 | 27.1 | 35.6 | 17.3 | 36.7 | 29.9 | 28.9 |
| 25-34 | 60.6 | 52.2 | 70.3 | 46.0 | 61.4 | 45.4 | 64.9 | 44.7 | 51.4 | 51.1 |
| 35-39 | 14.3 | 9.7 | 11.3 | 5.4 | 6.0 | 8.3 | 12.8 | 5.2 | 9.4 | 10.2 |
| ≥ 40 | 2.9 | 2.2 | 1.5 | † | † | † | 1.4 | 0.7 | 2.0 | 2.6 |
| Missing data | 0.0 | 0.0 | 0.0 | 0.0 | 0.0 | † | 0.0 | 0.0 | 0.0 | † |
| Maternal race/ethnicity |  |  |  |  |  |  |  |  |  |  |
| Non-Hispanic white | 90.1 | 86.3 | 94.0 | 91.1 | 91.2 | 91.1 | 93.5 | 91.8 | 94.2 | 82.8 |
| Non-Hispanic black | 4.6 | 8.0 | 0.9 | 3.7 | † | † | 1.4 | 2.4 | 1.6 | 11.2 |
| Hispanic | 1.7 | 2.0 | 1.3 | 1.6 | † | 3.0 | 1.5 | 2.9 | 1.3 | 2.1 |
| Asian/Pacific Islander | 1.1 | 1.8 | 3.2 | 3.0 | 4.8 | † | 2.8 | 1.9 | 2.3 | 1.4 |
| Native American | 1.9 | 1.5 | † | † | 0.0 | † | † | † | 0.6 | 2.0 |
| Non-Hispanic Other | 0.2 | 0.2 | 0.0 | † | † | 0.0 | 0.0 | † | † | 0.2 |
| Missing data | 0.4 | 0.3 | † | 0.0 | 0.0 | † | † | † | † | 0.4 |
| Maternal Education at Delivery |  |  |  |  |  |  |  |  |  |  |
| No High School Diploma | 6.8 | 12.4 | 19.0 | 18.3 | 17.1 | 13.1 | 13.9 | 19.0 | 16.7 | 9.7 |
| High School Diploma or equivalent | 26.5 | 42.7 | 47.3 | 51.8 | 51.0 | 44.2 | 48.2 | 49.9 | 54.1 | 38.6 |
| >High School Diploma | 66.2 | 44.6 | 33.3 | 29.6 | 31.5 | 40.0 | 37.5 | 31.1 | 29.1 | 51.4 |
| Missing Data | 0.5 | 0.4 | † | † | † | † | † | † | † | 0.4 |
| Maternal marital status |  |  |  |  |  |  |  |  |  |  |
| Married | 60.2 | 33.1 | 13.0 | 12.9 | 10.8 | 17.5 | 15.5 | 14.7 | 19.2 | 42.1 |
| Unmarried | 39.0 | 65.6 | 84.1 | 85.3 | 88.4 | 81.6 | 82.1 | 83.7 | 78.4 | 57.0 |
| Missing data | 0.8 | 1.3 | 2.9 | 1.8 | † | † | 2.4 | 1.5 | 2.4 | 0.9 |
| R-GINDEX† |  |  |  |  |  |  |  |  |  |  |
| Intensive | 11.1 | 12.3 | 14.3 | 7.8 | 10.8 | 7.7 | 15.3 | 10.6 | 13.3 | 12.3 |
| Adequate | 55.3 | 50.0 | 37.0 | 49.8 | 45.0 | 48.4 | 41.7 | 49.9 | 50.1 | 51.9 |
| Intermediate | 27.4 | 29.7 | 32.1 | 32.5 | 31.1 | 32.9 | 31.2 | 31.3 | 29.2 | 29.1 |
| Inadequate | 4.4 | 6.1 | 13.0 | 7.3 | 12.0 | 7.7 | 10.3 | 6.8 | 5.8 | 5.0 |
| None | 0.5 | 0.5 | 1.9 | † | † | † | † | † | 0.6 | 0.3 |
| Missing data | 1.4 | 1.3 | 1.6 | 1.8 | 0.0 | † | † | 1.1 | 1.0 | 1.3 |
| Plurality |  |  |  |  |  |  |  |  |  |  |
| Singleton birth | 96.6 | 96.7 | 96.3 | 97.8 | 92.4 | 98.8 | 96.2 | 97.2 | 96.5 | 96.7 |
| Plural Birth | 3.4 | 3.3 | 3.7 | 2.2 | 7.6 | † | 3.8 | 2.8 | 3.5 | 3.0 |
| Birth Weight, mean (SD) g | 3339.3 | 3275.3 | 3000.3 | 3089.5 | 3041.1 | 3231.1 | 3088.1 | 3170.7 | 3162.1 | 3356.7 |
| Low Birth Weight (<2500 g) |  |  |  |  |  |  |  |  |  |  |
| Yes | 7.3 | 9.1 | 18.9 | 15.3 | 17.9 | 11.0 | 14.8 | 11.0 | 12.1 | 6.6 |
| No | 92.7 | 90.9 | 81.0 | 84.7 | 82.1 | 89.0 | 85.2 | 89.0 | 87.8 | 93.3 |
| Missing data | 0.0 | 0.0 | † | 0.0 | 0.0 | 0.0 | 0.0 | 0.0 | † | † |
| Gestational age, mean, wks | 38.8 | 38.5 | 38.2 | 38.2 | 38.0 | 38.4 | 38.1 | 38.4 | 38.2 | 38.6 |
| Preterm birth (gestation <37 wks) |  |  |  |  |  |  |  |  |  |  |
| Yes | 8.8 | 10.1 | 15.0 | 14.7 | 19.1 | 10.7 | 14.6 | 10.5 | 11.3 | 8.9 |
| No | 91.0 | 89.8 | 84.8 | 85.3 | 80.9 | 89.3 | 85.4 | 89.5 | 88.6 | 91.1 |
| Missing data | 0.2 | 0.1 | † | 0.0 | 0.00 | 0.0 | 0.0 | 0.0 | † | 0.1 |
| Facility delivery volume§ |  |  |  |  |  |  |  |  |  |  |
| 10-249 | 18.0 | 23.0 | 19.6 | 19.4 | 20.7 | 16.6 | 16.7 | 24.9 | 30.4 | 22.4 |
| 250-499 | 7.5 | 7.7 | 5.5 | 3.9 | † | 3.3 | 7.5 | 14.1 | 7.4 | 7.5 |
| 500-999 | 25.1 | 25.4 | 18.6 | 26.6 | 29.5 | 36.8 | 17.4 | 23.7 | 22.7 | 26.8 |
| 1000-2499 | 23.0 | 24.9 | 36.9 | 37.6 | 31.9 | 27.0 | 30.0 | 23.9 | 24.7 | 22.9 |
| ≥2500 | 24.1 | 17.6 | 18.6 | 12.2 | 15.5 | 14.8 | 27.9 | 13.0 | 14.2 | 18.4 |
| Missing Data\|\| | 2.2 | 1.5 | 0.8 | † | † | † | † | † | 0.6 | 2.0 |

\*Other include sedative, hypnotic, or anxiolytic related disorders; other stimulant related disorders; hallucinogen related disorders; inhalant related disorders; and other psychoactive substance use related disorders.
†Data counts between 1 and 9 were suppressed for privacy reasons.
‡Measurement of the adequacy of prenatal care received during the prenatal period. Measure considers the number of prenatal visits, gestational age of the newborn, and the date of when prenatal care began.
§Average number of births per year.
||Missing data includes births that could not be categorized; homebirths (1222), en route to hospital, doctor's office/clinic or other (42), or were missing facility information (24).

Among the 3,228 Medicaid-linked birth records with drug dependency indicated on the birth record, 1,819 (56%) had at least one diagnosis for OUD in their medical claims during pregnancy, 821 (25%) cannabis use disorder (and no OUD nor cocaine use disorder), and 251 (8%) with alcohol, cocaine, nicotine, or other SUD (and no OUD nor cannabis use disorder) (**Table 3**, for non-mutually exclusive SUD diagnoses see **Supplemental Table 4**). No SUD diagnoses were found in the Medicaid claims for 337 (10%) of births where drug dependency was indicated on the birth record. Among the 24,220 Medicaid-linked birth records without drug dependency indicated on the birth record, 872 (4%) had at least diagnosis for OUD, 2,206 (9%) cannabis use disorder (and no OUD nor cocaine use disorder), and 3,086 (13%) with alcohol, cocaine, nicotine, or other SUD diagnosis (and no OUD nor cannabis use disorder) (**Table 3**). No SUD diagnoses were found in the Medicaid claims for 18,056 (75%) births.

Some of these patterns varied across the birth years. For example, among Medicaid-enrolled births with drug dependency indicated on the birth record, 63% had at least one OUD diagnosis in their Medicaid claims during pregnancy in 2016, but only 50% in 2020 (**Figure 1**). In 2016, cannabis use disorder diagnosis (and no OUD nor cocaine use disorder) was found in the Medicaid claims for 20% of births with drug dependency indicated on the birth record, which increased to 31% by 2020. Similar increases in cannabis use disorder during 2016-2020 were observed when we examined non-mutually exclusive SUD diagnoses (**Figure 2, Supplemental Table 4**) (which coincides with recreational marijuana use legalization in Maine in 2016).^20^

**Figure 1.**
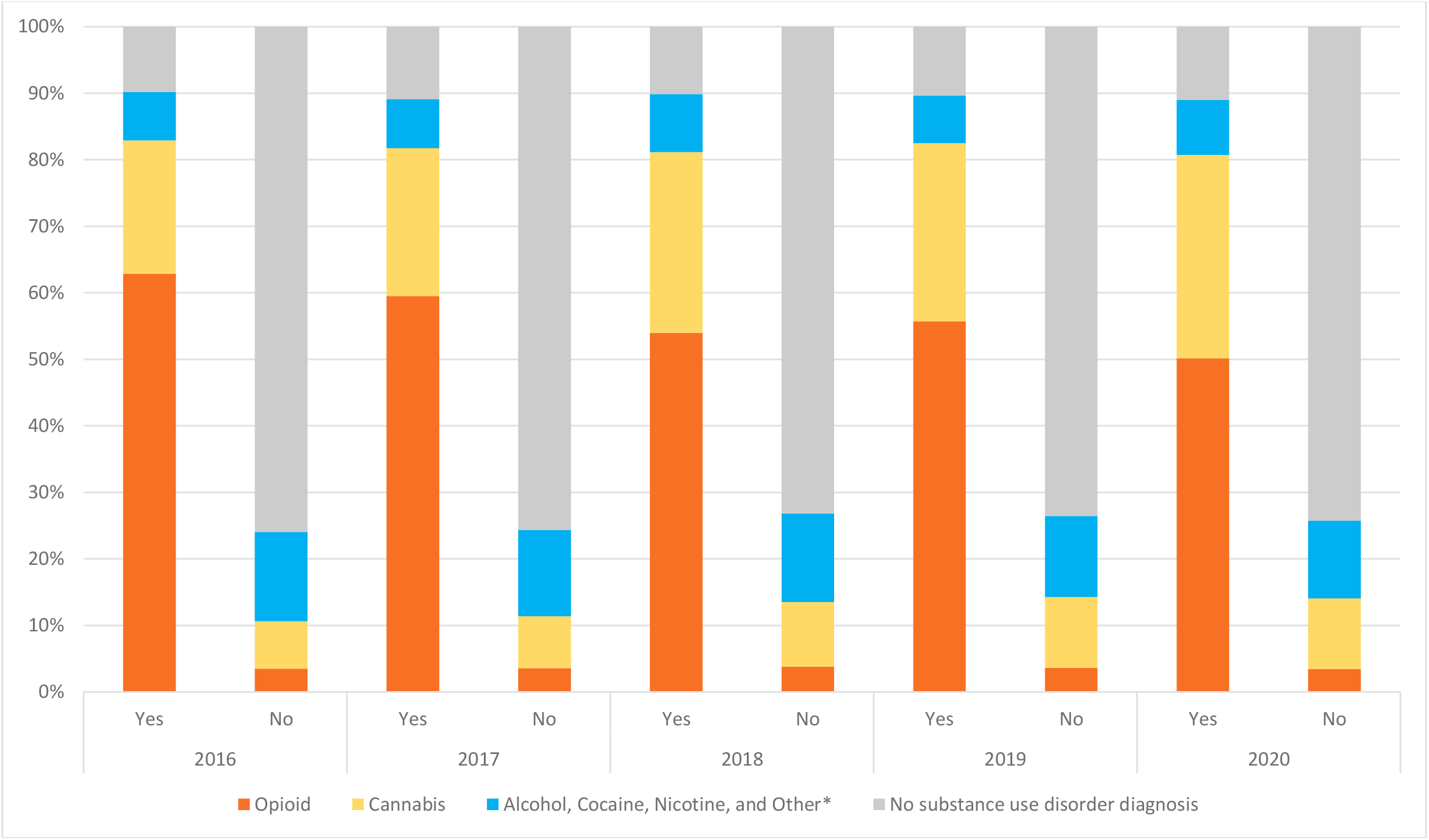
Maine birth records for in-state resident livebirths among women Medicaid enrolled at month of delivery by drug dependency checkbox and hierarchical categorization of substance use disorder in medical claims during pregnancy, 2016-2020 (n=27,448) *Other SUD diagnoses include sedative, hypnotic, or anxiolytic related disorders; other stimulant related disorders; hallucinogen related disorders; inhalant related disorders; and other psychoactive substance use related disorders.

**Figure 2.**
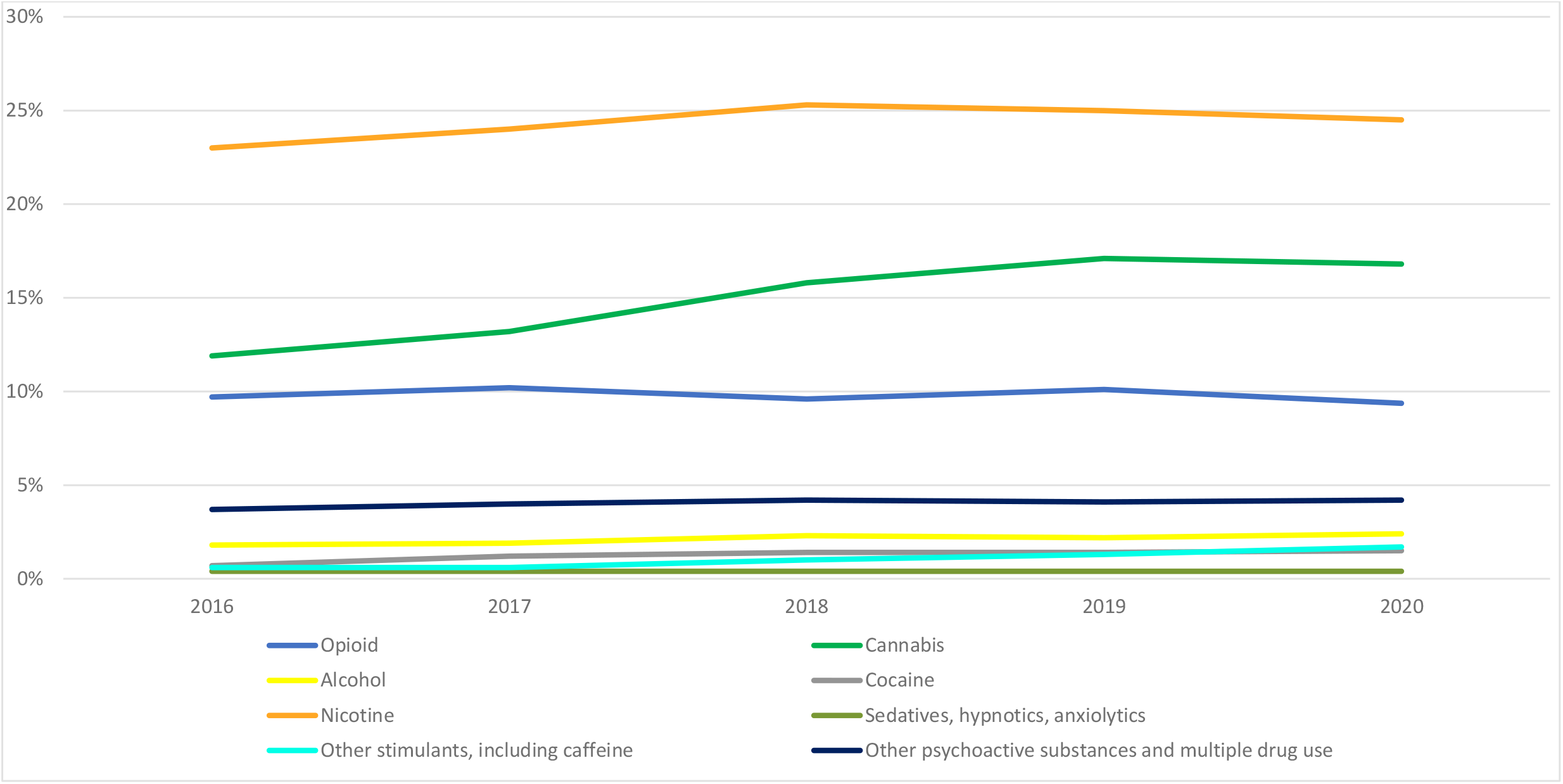
Maine women Medicaid enrolled at month of delivery by non-mutually exclusive drug-specific substance use disorder* in medical claims during pregnancy, 2016-2020 (n=27,448) **\***These included mental and behavioral disorders due to use of alcohol (ICD-10-CM F10.xxx), use of opioids (F11.xxx), use of cannabis (F12.xxx), use of sedatives, hypnotics, anxiolytics (F13.xxx), use of cocaine (F14.xxx), use of other stimulants, including caffeine (F15.xxx), use of hallucinogens (F16.xxx), use of nicotine (F17.xxx), use of inhalants (F18.xxx), use of other psychoactive substances and multiple drug use (F19.xxx). Substance use disorders shown in this figure are not mutually exclusive. Disorders due to use of hallucinogens and inhalants suppressed from presentation because counts between 1 and 9 per year.

Among the births to women who were enrolled in Medicaid at the month of delivery with drug dependency indicated on the birth record and at least one OUD diagnosis in their Medicaid claims, 81% had MOUD documented in their medical claims during pregnancy (**Supplemental Table 5**). Among women without drug dependency indicated on the birth record, 48% had MOUD documented during pregnancy.

In our sensitivity analysis restricted to women enrolled in Medicaid for their entire pregnancy, 15% had drug dependency indicated on their birth record and 41% had at least one SUD diagnosis in their Medicaid claims data, which was higher than in our primary analysis (12% and 33%, respectively) (**Supplemental Table 6**).

## DISCUSSION

Among 58,584 births in Maine during 2016-2020, we found 47% (n=27,448) were enrolled in Medicaid at the month of delivery. Drug dependency as indicated by a checkbox on the birth record was higher among women with Medicaid listed as their primary payer compared to all other payers except Medicare. While diagnoses of OUD and cannabis use disorder (without OUD or cocaine use disorder) during pregnancy in Medicaid claims data were higher among those with drug dependency indicated on the birth record, 1 in 10 birth records with the checkbox checked had no evidence of SUD during pregnancy and 1 in 4 birth records with the checkbox unchecked had evidence of SUD during pregnancy. We conclude that reporting of drug dependency on the birth record does not appear to accurately indicate SUD during pregnancy. Our findings suggest the drug dependency checkbox on the Maine birth certificate may be of limited value in identifying SUD during pregnancy.

Prior research on the accuracy of the birth record to assess maternal SUD is limited. A study analyzing Oregon births over a one-month period in 1989 found that birth certificate reporting identified only 41% of women with recognized illicit drug use.^21^ While relevant to our findings, this study was conducted nearly 30 years ago, before the current opioid epidemic in the US began. Several other studies have analyzed the accuracy of birth certificate data compared to medical records or Medicaid data for tobacco and alcohol use, finding generally low agreement between the data sources.^22-26^ Additionally, the Iowa Health in Pregnancy Study compared birth certificate and maternal recall data from livebirths during 2002-2005, also finding low agreement between the data sources for smoking and alcohol use during pregnancy.^27^ One explanation for these discrepancies in SUD documentation is a hesitancy to report health-limiting behaviors during pregnancy to medical providers and birth certifier for fear of repercussions.^27^

Further, the way the birth record is completed may also influence its accuracy. Although there are detailed instructions for how each item on the US Standard Certificate of Live Birth should be completed,^12,13^ as a state-specific item, the drug checkbox was not accompanied by explicit instructions for completion. Therefore, lack of instructions for which drugs and frequency of drug use count as “drug dependency", what time frames are considered (e.g. during the entire pregnancy or at the time of delivery), and how treatment for SUD and legal status of the drug being used should be handled, likely have led to variation in how the drug dependency checkbox is used. Our study found that women with OUD who had drug dependency indicated on their birth record were more likely to be in MOUD treatment as compared with women with OUD who did not have drug dependency indicated on their birth certificate. This suggest that women in treatment might be more noticeable to their provider as having SUD, and therefore more likely to be recorded as such on their birth record.

### Strengths and limitations

Our study had several strengths. First, we examined SUD diagnoses among women covered by Medicaid during pregnancy, who account for more than 80% of women with OUD during pregnancy nationally and in Maine.^2,28^ Second, Maine Medicaid/CHIP (MaineCare) has used a fee-for-service model to pay providers since the late 1990’s; this means that claims are collected and submitted in a standardized way for each service provided, an advantage over other states that use managed care arrangements.^29^

However, our study had some limitations. The number of women with cocaine and alcohol use disorders was small and therefore collapsed with other SUD into a single category, resulting in a heterogenous group largely comprised of those with nicotine use disorder. Additionally, because there was no definition for what the drug dependency checkbox indicated, we did not know how to define an appropriate comparison using the Medicaid data, e.g. what drug disorders to include, perinatal timeframe to examine, and how to classify women in SUD treatment. Finally, Maine modified the drug dependency item on the birth record in March 2021 to separately capture “alcohol use disorder” and “substance use disorder” (**Supplemental Figure 1**). This item will be further changed in late 2022 to capture specific drugs used during pregnancy, which will be shared with the Office of Child and Family Services to help identify substance exposed infants (personal communication, Director of DRVS). This means our analysis may not be relevant to Maine birth records going forward.

In conclusion, although diagnoses of OUD and cannabis use disorder were more common among Maine birth records with drug dependency indicated, reporting of drug dependency on the birth record does not accurately indicate SUD during pregnancy. Our findings suggest the drug dependency checkbox on the Maine birth certificate may be of limited value in identifying SUD during pregnancy, and that plans to separately capture specific drugs used during pregnancy appear warranted. Explicit instructions as to how to complete this item may improve its accuracy.

## Data Availability

All data produced in the present study are available upon reasonable request to the authors

## ACKNOWLEDGEMENTS

We thank the following employees of the Maine Department of Health and Human Services for supporting this project and providing comments that greatly improved the manuscript: Kim Haggan, Director of Data, Research, and Vital Statistics; Liz Remillard, Program Manager, MaineMOM Initiative; David Jorgenson, Director of Data Analytics, Office of MaineCare Services; Oliva Alford, Director, Delivery System Reform, Office of MaineCare Services; and Michelle Probert, Medicaid Director, Office of MaineCare Services. We are also immensely grateful to Erika Lichter and Fleur Hopper, of the Maine Center for Disease Control, for allowing us to use their SAS code to import the birth certificate text files.

---

**Supplemental Table 1:** Linkage rules for maternal Medicaid enrollee linkage to the birth record, Maine, Jan 2016-Dec 2020

| Weight | Points of linkage | Rules |
| --- | --- | --- |
| #1 | 3 | Full DOB, Name, Medicaid ID |
| #2 | 3 | Full DOB, Name, 4 Digits for SSN |
| #3 | 3 | Full DOB, 4 Digits for SSN, Medicaid ID |
| #4 | 3 | DOB = Month & Year only, Name, Medicaid ID |
| #5 | 3 | DOB = Month & Year only, Name, 4 Digits for SSN |
| #6 | 3 | Full DOB, First Name First Letter, Last Name = last Name or Last Name = Maiden or Last Name of mom = child's last name, Medicaid ID |
| #7 | 3 | Full DOB, First Name First Letter, Last Name = last Name or Last Name = Maiden or Last Name of mom = child's last name, 4 Digits for SSN |
| #8 | 3 | Full DOB, Last Name = last Name or Last Name = Maiden or Last Name of mom = child's last name, 4 Digits of SSN |
| #9 | 3 | Full DOB, First Name, Middle Initial and Medicaid ID |
| #10 | 3 | Full DOB, First Name, Middle Initial and 4 Digits for SSN |
| #11 | 2 | Last Name = last Name or Last Name = Maiden or Last Name of mom = child's last name, Medicaid ID, 4 Digits for SSN |
| #12 | 2 | Full DOB, Name |
| #20 | varies | Manually check of possible linkages (said Medicaid would pay for delivery or reported Medicaid ID, but didn't link) based on date of delivery |
| #30 | varies | Manual maternal linkages based on infant linkages |

\*"Name" means: First Name= First Name AND (Last Name =Last Name OR Last Name = Maiden Name OR Last Name of mom = child's last name)
†DOB= Date of birth
‡SSN= Social Security Number

**Supplemental Table 2:**
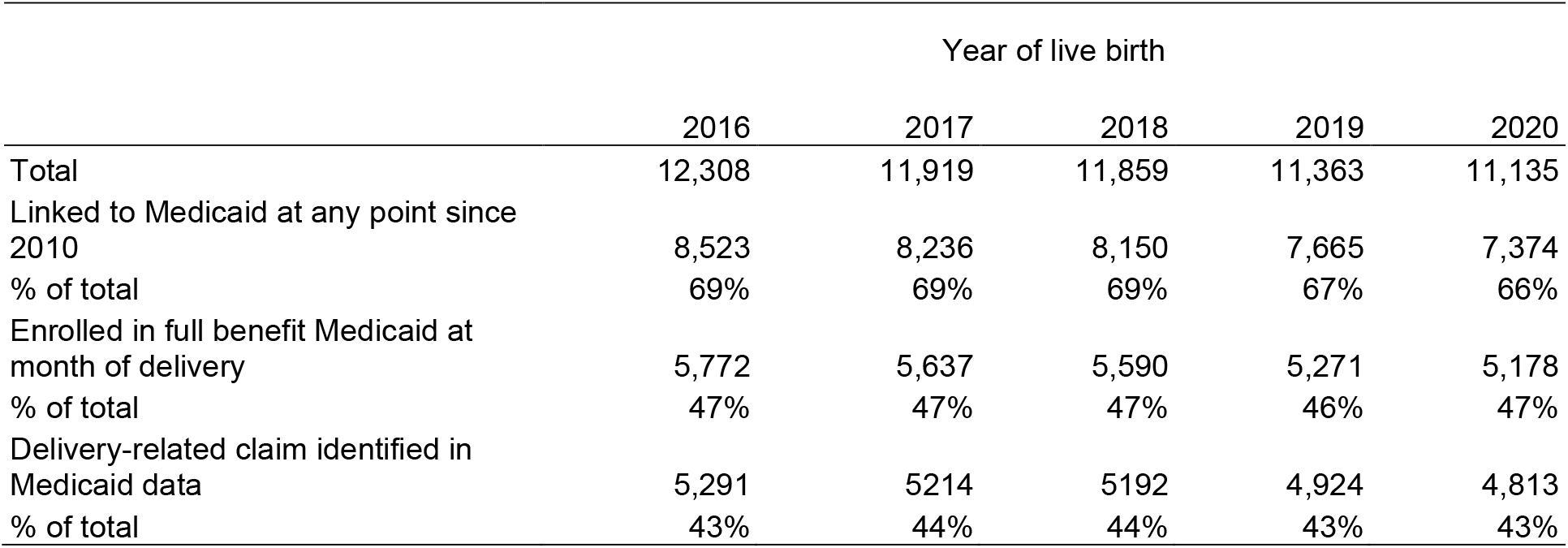
Results for maternal Medicaid enrollee linkage to the birth record, Maine, Jan 2016-Dec 2020, N=58,584

**Supplemental Table 3:** Rule-based deterministic linkage results for maternal Medicaid enrollee linkage to the birth record, Maine, Jan 2016-Dec 2020, N=58,584

|  | 2016 | 2017 | 2018 | 2019 | 2020 | Total |
| --- | --- | --- | --- | --- | --- | --- |
| Weight |  |  |  |  |  |  |
| No match | 3785 | 3683 | 3709 | 3698 | 3761 | 18636 (32%) |
| #1 | 4000 | 3834 | 3903 | 3549 | 3639 | 18925 (32%) |
| #2 | 3831 | 3732 | 3638 | 3536 | 3187 | 17924 (31%) |
| #3 | 239 | 205 | 158 | 173 | 175 | 950 (2%) |
| #4 | 7 | 14 | 19 | 8 | 4 | 52 (0%) |
| #5 | 10 | 7 | 10 | 8 | 14 | 49 (0%) |
| #6 | 7 | 5 | 7 | 11 | 12 | 42 (0%) |
| #7 | 67 | 64 | 54 | 53 | 49 | 287 (0%) |
| #8 | 4 | 2 | 6 | 3 | 4 | 19 (0%) |
| #9 | 3 | 6 | 3 | 3 | 2 | 17 (0%) |
| #10 | 103 | 96 | 77 | 78 | 78 | 515 (1%) |
| #11 | 14 | 18 | 8 | 8 | 10 | 58 (0%) |
| #12 | 187 | 194 | 213 | 165 | 124 | 883 (2%) |
| #20 | 15 | 19 | 20 | 28 | 37 | 119 (0%) |
| #30 | 36 | 40 | 34 | 42 | 39 | 191 (0%) |
| Total | 12308 | 11919 | 11859 | 11363 | 11135 | 58584 (100%) |

\*See Supplemental Table 1 for explanation of rules.

**Supplemental Table 4.** Maine birth records for in-state resident livebirths among women Medicaid enrolled at month of delivery by drug dependency checkbox and specific substance use disorder in medical claims during pregnancy, 2016-2020 (n=27,448)

| Substance use disorder* | Drug Dependence = Yes on Birth Certificate | Drug Dependence = No on Birth Certificate |
| --- | --- | --- |
| Opioid | 1819 (56.4) | 872 (3.6) |
| Cannabis | 1575 (48.8) | 2507 (10.4) |
| Alcohol | 210 (6.5) | 372 (1.5) |
| Cocaine | 232 (7.2) | 103 (0.4) |
| Nicotine | 2130 (66.0) | 4552 (18.8) |
| Sedatives, hypnotics, anxiolytics | 60 (1.9) | 46 (0.2) |
| Other stimulants, including caffeine | 154 (4.8) | 117 (0.5) |
| Hallucinogen | † | † |
| Inhalant | 16 (0.5) | † |
| Other psychoactive substances and multiple drug use | 689 (21.3) | 417 (1.7) |

\*These included mental and behavioral disorders due to use of alcohol (ICD-10-CM F10.xxx), use of opioids (F11.xxx), use of cannabis (F12.xxx), use of sedatives, hypnotics, anxiolytics (F13.xxx), use of cocaine (F14.xxx), use of other stimulants, including caffeine (F15.xxx), use of hallucinogens (F16.xxx), use of nicotine (F17.xxx), use of inhalants (F18.xxx), use of other psychoactive substances and multiple drug use (F19.xxx). Substance use disorders shown in this table are not mutually exclusive.
†Data counts between 1 and 9 were suppressed for privacy reasons.

**Supplemental Table 5.** Medication for opioid use disorder in Medicaid claims for 2,691 Maine births among women enrolled in Medicaid at the month of delivery with opioid use disorder, 2016-2020

|  | Drug Dependence = Yes on Birth Certificate |  |  |  |  |  | Drug Dependence = No on Birth Certificate |  |  |  |  |  |
| --- | --- | --- | --- | --- | --- | --- | --- | --- | --- | --- | --- | --- |
| **MOUD\* in Medicaid claims** | Opioid use disorder documented in Medicaid Claims |  |  |  |  |  | Opioid use disorder documented in Medicaid Claims |  |  |  |  |  |
|  | 2016-2020<br>(n= 1,819) | 2016<br>(n=379) | 2017<br>(n=394) | 2018<br>(n=352) | 2019<br>(n=366) | 2020<br>(n=328) | 2016-2020<br>(n= 872) | 2016<br>(n=182) | 2017<br>(n=179) | 2018<br>(n=187) | 2019<br>(n=167) | 2020<br>(n=157) |
| MOUD Procedure | 394 (21.7) | 101 (26.7) | 97 (24.6) | 62 (17.6) | 70 (19.1) | 64 (19.5) | 94 (10.8) | 28 (15.4) | 17 (9.5) | 17 (9.1) | 14 (8.4) | 18 (11.5) |
| MOUD Prescription | 1,175 (64.6) | 244 (64.4) | 262 (66.5) | 238 (67.6) | 227 (62.0) | 204 (62.2) | 346 (39.7) | 72 (39.6) | 74 (41.3) | 74 (39.6) | 63 (37.7) | 63 (40.1) |
| Either MOUD procedure or prescription | 1,471 (81.0) | 322 (85.0) | 333 (84.5) | 280 (79.5) | 284 (77.6) | 252 (76.8) | 421 (48.3) | 97 (53.3) | 88 (49.2) | 88 (47.1) | 72 (43.1) | 76 (48.4) |

\*MOUD = Medication for opioid use disorder

**Supplemental Table 6.** Medicaid linkage to Maine birth records by drug dependency checkbox on birth record, among women enrolled in full-benefit Medicaid for entire pregnancy and not dual enrolled in Medicare or with a private payer, 2016-2020

| Primary Payer<br>Reported on Birth<br>Record | Medicaid<br>Enrolled at<br>Month of<br>Delivery, N | Medicaid Enrolled<br>for Entire<br>Pregnancy, N | Drug Dependency<br>on Birth Record |  | Any Substance Use Disorder Diagnosis in<br>Medicaid Claims* |  |
| --- | --- | --- | --- | --- | --- | --- |
|  |  |  | Among Medicaid Enrolled for Entire<br>Pregnancy |  | Among Medicaid Enrolled for Entire Pregnancy |  |
|  |  |  | Yes,<br>n (%) | No,<br>n (%) | Yes,<br>n (%) | No,<br>n (%) |
| Medicaid | 22,837 | 8,932 | 1,326 (14.9) | 7,606 (85.2) | 3,756 (42.1) | 5,176 (58.0) |
| Private Insurance | 3,426 | 178 | † | 169 (94.9) | 30 (16.9) | 148 (83.2) |
| Self-Pay | 862 | 141 | 16 (11.4) | 125 (88.7) | 38 (30.0) | 103 (73.1) |
| Indian Health Service | 14 | † | † | † | † | † |
| Champus/Tricare | 64 | † | † | † | † | † |
| Other Government | 67 | † | † | † | † | † |
| Other/Unknown | 178 | 22 | † | 21 (95.5) | † | 16 (72.7) |
| Total | 27,488 | 9,288 | 1354 (14.6) | 7934 (85.4) | 3835 (41.3) | 5,453 (58.7) |

\*Substance use disorders include alcohol related disorders; opioid related disorders; cannabis related disorders; sedative, hypnotic, or anxiolytic related disorders; cocaine related disorders; other stimulant related disorders; hallucinogen related disorders; nicotine dependence; inhalant related disorders; and other psychoactive substance use related disorders.
†Counts between 1 and 9 were suppressed for privacy purposes.

**Supplemental Figure 1:**
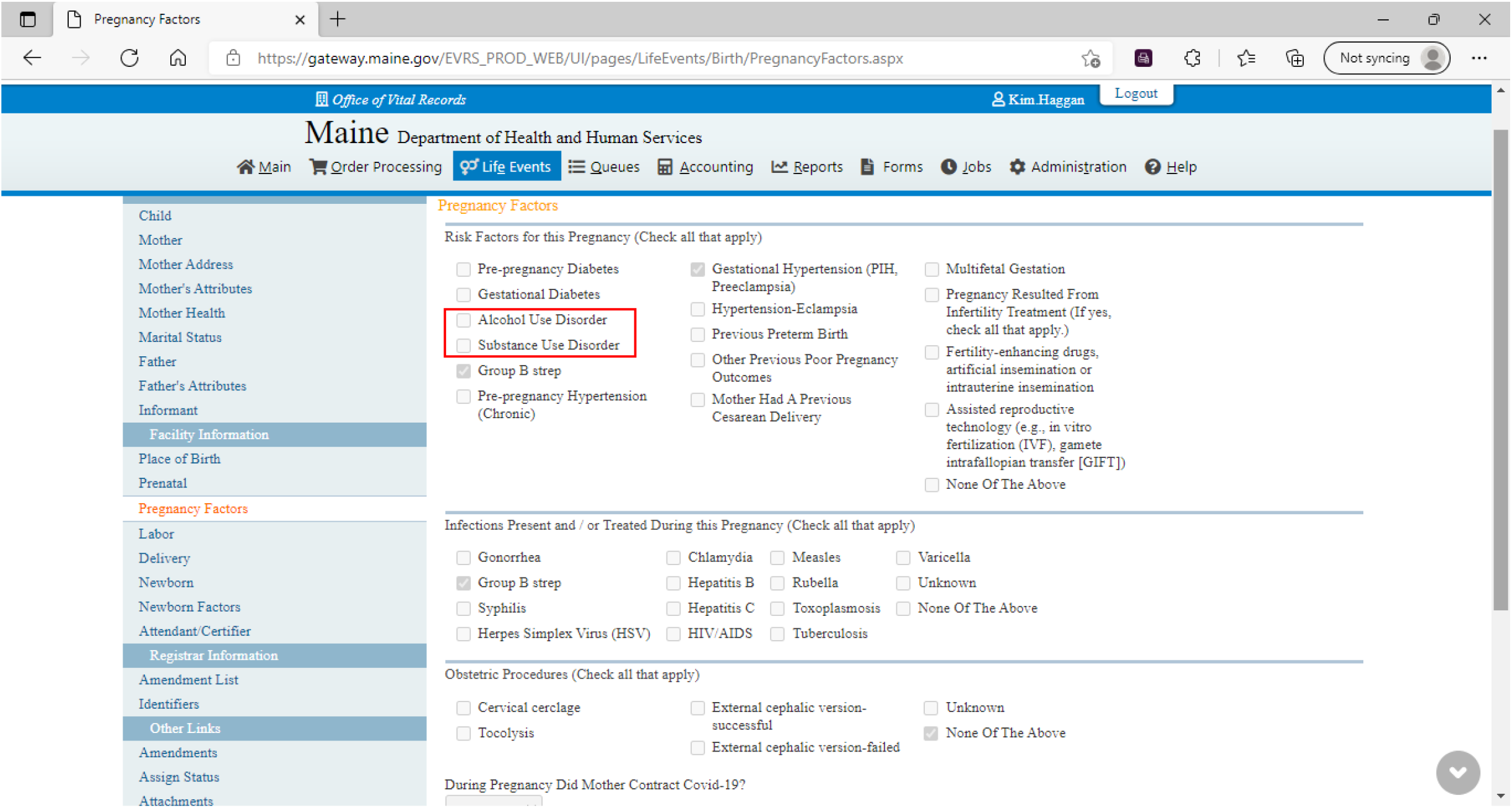
Screenshot of electronic entry screen for Maine birth records “Pregnancy Factors", which contains the 2021 revised drug dependency checkboxes for Alcohol Use Disorder and Substance Use Disorder. These checkboxes will be further revised in late 2022.

